# The association between immunosuppressants use and COVID-19 adverse outcomes: National COVID-19 cohort in South Korea

**DOI:** 10.1101/2021.08.17.21262183

**Authors:** Hyun Joon Shin, Ronald Chow, Hyerim Noh, Jongseong Lee, Jihui Lee, Young-Geun Choi

## Abstract

**Purpose:** There is uncertainty of the effect of immunosuppression, including corticosteroids, before COVID-19 infection on COVID-19 outcomes. The aim of this study was to investigate the relationship between prehospitalization immunosuppressants use (exposure), and COVID-19 patient outcomes.

**Methods:** We conducted a population-based retrospective cohort study using a nationwide healthcare claims database of South Korea as of May 15, 2020. Confirmed COVID-19 infection in hospitalized individuals aged 40 years or older were included for analysis. We defined exposure variable by using inpatient and outpatient prescription records of immunosuppressants from the database. Our primary outcome was a composite endpoint of all-cause death, intensive care unit (ICU) admission, and mechanical ventilation use. Inverse probability of treatment weighting (IPTW)-adjusted logistic regression analyses were used, to estimate odds ratio (OR) and 95% confidence intervals, comparing immunosuppressants users and non-users.

**Results:** We identified 4,349 patients, for which 1,356 were immunosuppressants users and 2,903 were non-users. Patients who used immunosuppressants were at increased odds of the primary outcome of all-cause death, ICU admission and mechanical ventilation use (IPTW OR 1.32; 95% CI: 1.06 – 1.63). Patients who used corticosteroids were at increased odds of the primary outcome (IPTW OR 1.33; 95% CI: 1.07 – 1.64).

**Conclusion:** We support the latest guidelines from the CDC, that people on immunosuppressants are at high risk of severe COVID-19 and immunocompromised people may need booster COVID-19 vaccinations.

**Funding:** YGC’s work was partially supported by 2020R1G1A1A01006229 awarded by the National Research Foundation of Korea.

## Introduction

In December 2019, an outbreak of a novel coronavirus was reported in Wuhan, China, which was later named the severe acute respiratory syndrome coronavirus 2 (SARS-CoV-2) by the Coronavirus Study Group of the International Committee on Taxonomy of Viruses^1,2^. The SARS-Cov-2 virus rapidly spread around the world, causing the COVID-19 disease in infected patients; on March 12, 2020, the World Health Organization (WHO) officially declared COVID-19 a pandemic^3^.

As the scientific community raced to find possible active agents for treatment and cure of COVID-19, supportive care therapies were developed and tested^3,4^. Immunosuppressants, specifically corticosteroids, were employed, as it could help COVID-19 patients by mitigating cytokine storms^5^. But, there are concerns that it can worsen viral shedding, and increase mortality due to COVID-19^6^.

The current WHO clinical guidelines, as of November 20, 2020, provides a strong recommendation for corticosteroids use in severe and critical patients with COVID-19 disease^7^. However, in patients with mild to moderate COVID-19 infection who do not require oxygen support, corticosteroids use was associated with a trend towards higher mortality, although not statistically significant in the RECOVERY trial (17.8% vs 14%; RR 1.19 and 95% CI: 0.92-1.55)^8^. As a result, there is uncertainty of the effect of immunosuppression, including corticosteroids started before COVID-19 infection, on COVID-19 outcomes since patients on immunosuppression could have any spectrum of COVID-19 disease, from asymptomatic to critical COVID-19 infection. It is possible that patients with non-severe COVID-19 infection could receive harm from baseline corticosteroids use.

The aim of this study was to investigate the relationship between immunosuppressants use, including corticosteroids, and COVID-19 patient outcomes.

## Methods

### Data Source

The Health Insurance Review and Assessment Service (HIRA) of South Korea is the sole nationwide government agency that operates a fee-for-service reimbursement system, which covers 98% of the Korean population. Its administrative claims database includes the beneficiary’s sociodemographic characteristics, healthcare utilization history, diagnosis results (International Classification of Diseases, 10^th^ Revision), as well as prescription from both inpatient and outpatient settings.

On March 27, 2020, the #OpenData4COVID19 project by the Ministry of Health and Welfare (MOHW) of Korea released a patient-level, deidentified COVID-19 data based on the HIRA insurance claims database, which is the first nationwide dataset of COVID-19 patients. The HIRA COVID-19 database included data for all individuals who received a reverse transcription-polymerase chain reaction (RT-PCR) test for COVID-19 as of May 15, 2020, which was linked to their claims data for the previous three years.

This study was approved by the Human Investigation Review Board of Public Institutional Bioethics Committee designated by the MOHW, which waived the requirement of informed consent due to retrospective study design and anonymity of the HIRA database (IRB # P01-2020-1262-001.

### Study Design and Participants

The HIRA data consisted of 234,427 individuals who were tested for SARS-CoV-2 between January 1, 2020 and May 15, 2020. 7,590 were identified positive for COVID-19, as designated by the coding in the #OpenData4COVID19 project (Supplementary Material 2). 4,610 individuals were aged 40 years or older. We excluded those who were less than 40 years old, as it is unlikely that these younger individuals would experience severe COVID-19 outcomes. For precise outcome measurements, we further restricted to individuals who were hospitalized for COVID-19, resulting in 4,349 individuals in our study cohort (Figure 1). The cohort entry date was defined as the admission date of COVID-19 hospitalization. In South Korea, individuals who were confirmed COVID-19 positive were required hospitalization until full recovery, defined by the cessation of fever without medication use and two negative test results within 24 hours^9^. However, there were a small number of confirmed COVID-19 patients who were not hospitalized due to the temporal shortage of health facilities.

**Figure 1.**
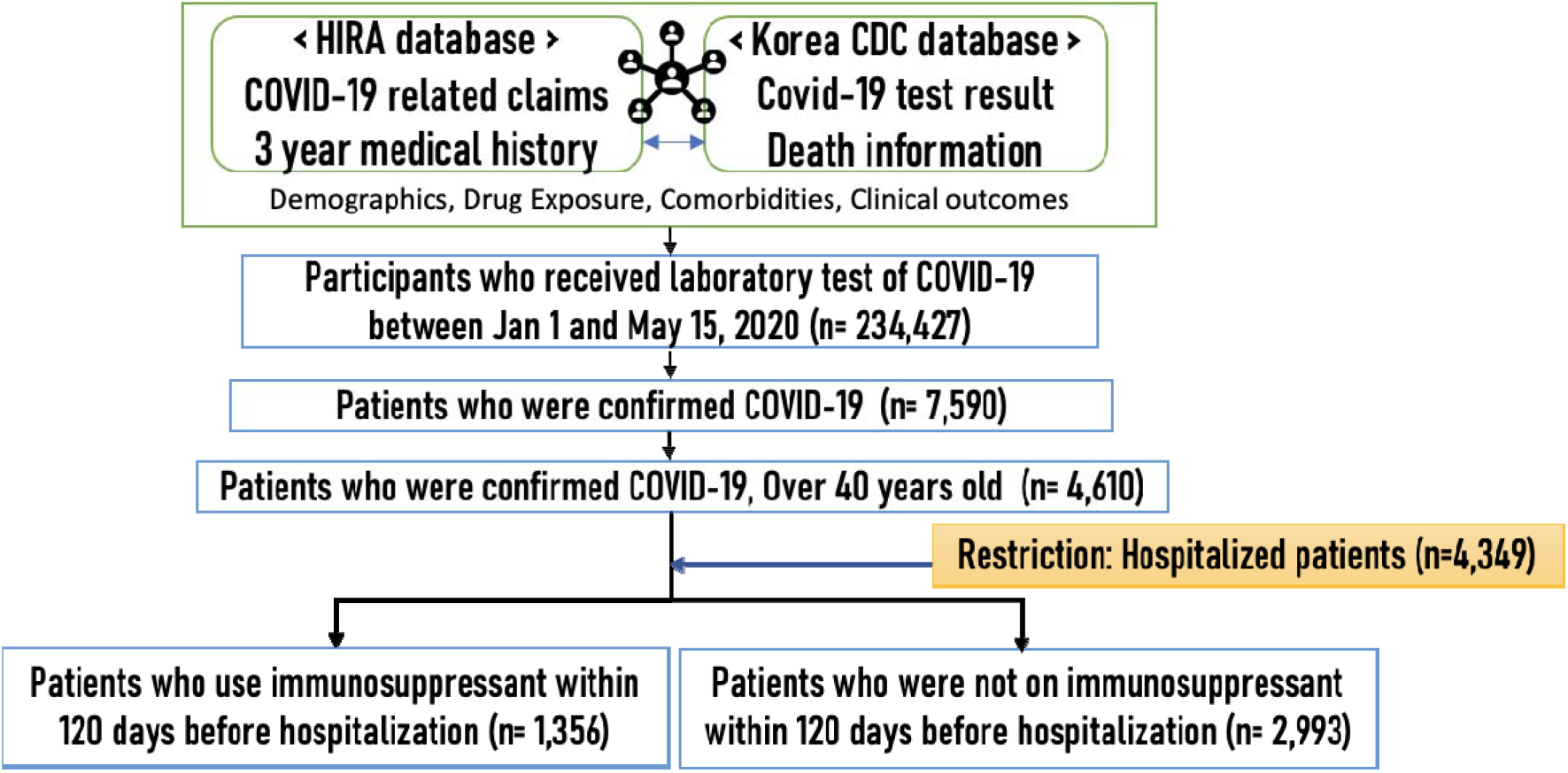
Nationwide population-based cohort study design using the HIRA and KDCA database of South Korea for the entire South Korean population of 50 million inhabitants. Hospitalized patients aged >= 40 who used immunosuppressants within 120 days before COVID-19 test were classified as immunosuppressants users while patients who were not on immunosuppressants within 120 days were classified as non-users. Abbreviations: COVID-19, coronavirus 2019; HIRA, Health Insurance Review and Assessment Service; KDCA, Korean Disease Control and prevention Agency.

### Immunosuppressant Exposure

We classified individuals who had inpatient and outpatient prescription records of immunosuppressants at least once within 120 days prior to the cohort entry (ascertainment window: -120d to 0d) as immunosuppressants users. Other individuals were defined as non-users. Among immunosuppressants users, corticosteroids users were assessed as a separate exposure group and compared to the non-immunosuppressants-users in sensitivity analysis. Our definition follows an intention-to-treat approach. Immunosuppressants were identified by the anatomical therapeutic chemical (ATC) codes (Supplementary Material 2).

### Outcomes

Our primary outcome was a composite endpoint of all-cause death, intensive care unit (ICU) admission, and mechanical ventilation use. As secondary outcomes, we evaluated the individual components, of the primary composite endpoint. In-hospital ICD-10 diagnostic codes and the national procedure codes were used to define the outcomes (Supplementary Material 2). We measured these study outcomes between the cohort entry date through the end of follow-up (discharge, or end of study).

### Potential Confounders

We considered sociodemographic and clinical factors as potential confounders. We included sex, age (in years), age square and health insurance type at cohort entry for sociodemographic factors. Clinical factors consisted of 20 comorbidities and 12 co-medications uses. The complete list of clinical factors and their working definitions is described in Supplementary Material 2. We defined comorbidity variables using in-hospital ICD-10 diagnostic codes, with an index period from 3 years prior (−1080d) through the start of the evaluation of exposure (−120d). When defining malignancy, we additionally employed the expanded benefit coverage codes, to minimize false-positive classification. For co-medication use, we used in/out-hospital ATC codes with an index period from -240d to -120d.

### Statistical Analyses

We described baseline characteristics for immunosuppressants users and non-users as counts with percentage (for categorical variables) and means with standard deviations (for continuous variables). We summarized distributional imbalances between two groups by absolute standardized difference (aSD) for each variable. Empirically, aSD ≤ 0.1 is preferred for balance between groups.

For outcome analysis, we employed the inverse probability of treatment weight (IPTW) approach^10,11^, based on propensity scores (PS), the probability for an individual to receive immunosuppressants. We estimated PS by a multivariable logistic regression that included all confounders in Table 1 as predictors. Every individual was weighted by either 1/PS (the immunosuppressants users) or 1/(1-PS) (non-users). This led to a balanced pseudo-population. Then, we fitted weighted univariate logistic regression models to estimate odds ratio (OR) with 95% confidence intervals (CI) for the association effect of immunosuppressants on each outcome. In addition, for comparison purposes, we fitted unweighted univariate logistic regression models and unweighted multivariate logistic regression models, adjusting for age, sex, insurance type, history of diabetes and history of hypertension.

**Table 1.**
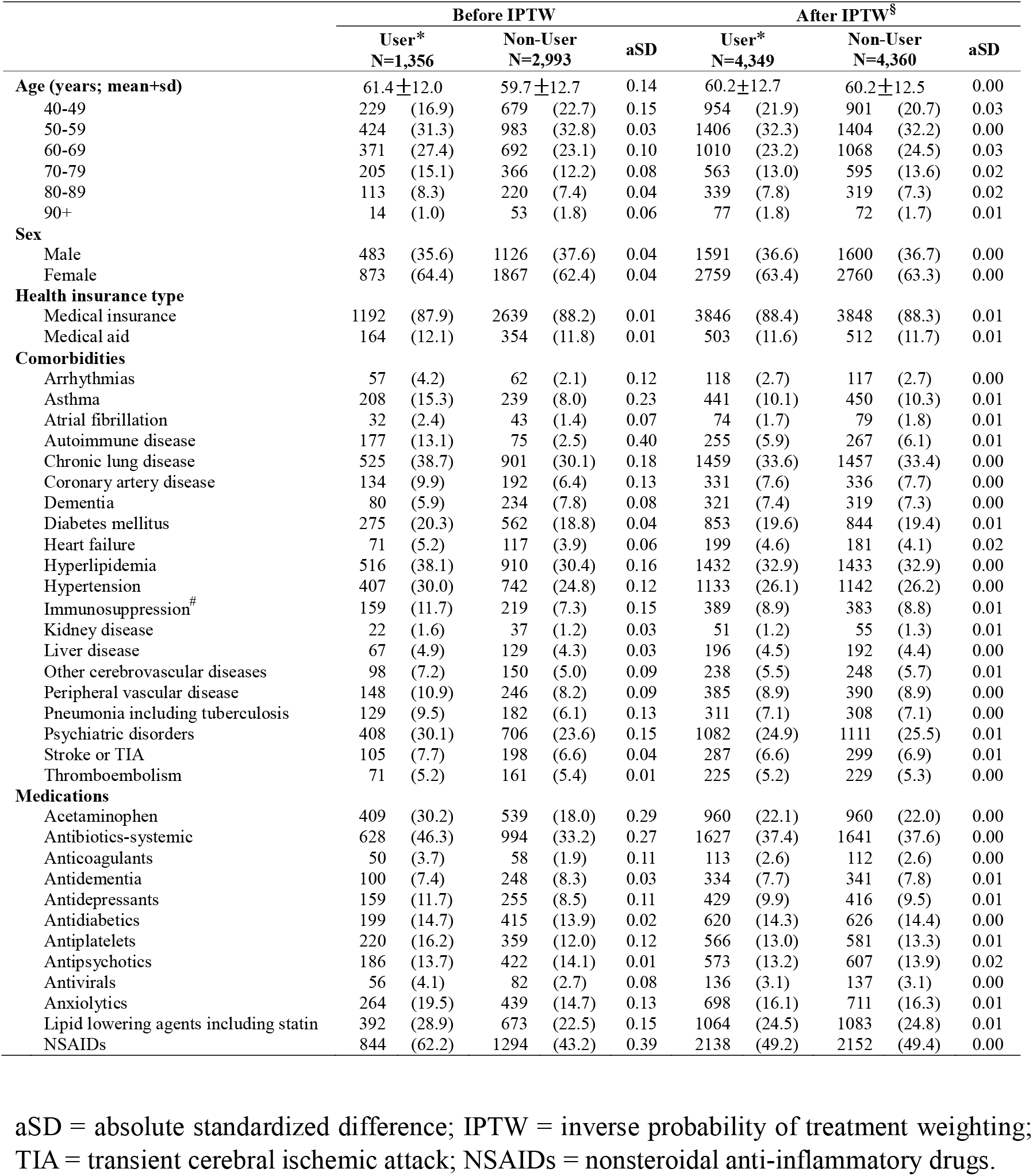
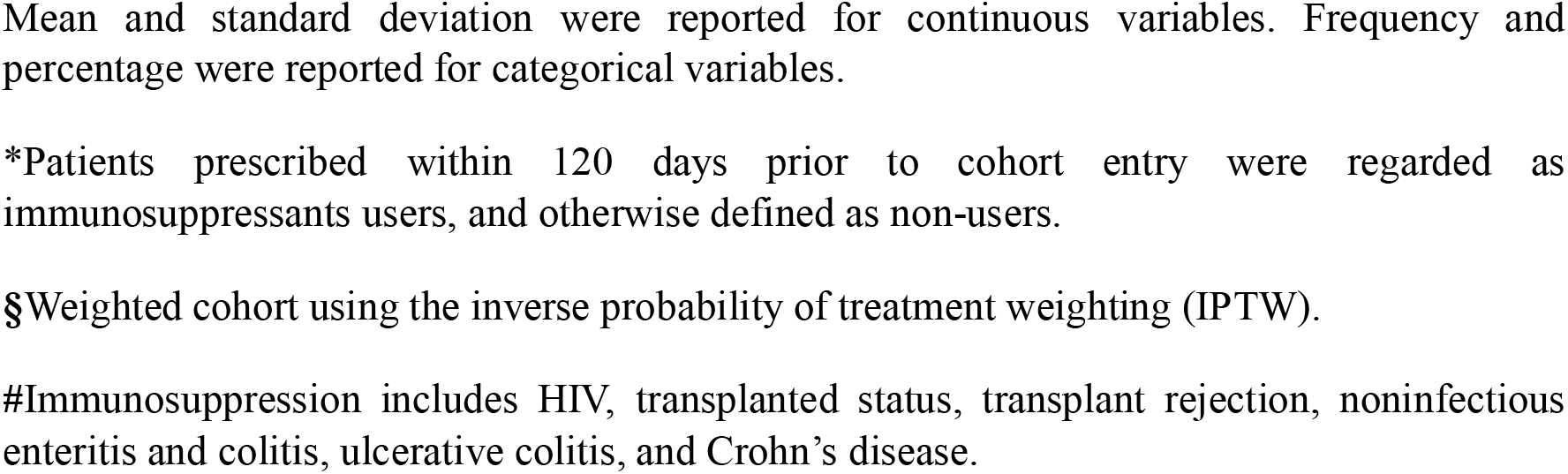
Baseline sociodemographic and clinical characteristics of adult patients(>=40years old) with COVID-19 in South Korea, as of May 15, 2020 (Users: Immunosuppressant within 120days)

### Statistical Analyses – Subgroup Analyses

We conducted a subgroup analysis for the risk of the primary outcome. We considered stratifications by the following: (1) sex; (2) age, classified into groups with age < 65 years and ≥ 65 years; (3) the history of autoimmune disease, cancer and HIV, and (4) the history of autoimmune disease. For each subgroup analysis, we conducted a multivariate IPTW-weighted logistic regression that includes the exposure (immunosuppressants use), stratification variable and the interaction of those two. We obtained the p-value of the interaction term (p for interaction). For the weight used in subgroup analyses, we recalculated PS using multivariate logistic regression.

### Statistical Analyses – Sensitivity Analyses

We investigated the sensitivity of results against the scope of working definitions. First, we redefined the study population by those who were aged ≥ 40 years and confirmed positive, which relaxes the restriction of hospitalization. Second, to exclude individuals who used immunosuppressants for a short period but stopped prior to hospitalization, we narrowed down the window of ascertaining exposure, from 120 days to 90 days. Third, we narrowed down the definition of the exposure group from immunosuppressants users to corticosteroids users. For each change of the settings, we repeated our main analysis.

To examine the sensitivity of results against the selection of statistical tools, the following alternative approaches were considered. First, we excluded subjects with extreme PS values, that is, PS < 0.025 or PS > 0.975 (IPTW with trimming). Second, we additionally included the fitted PSs to other covariates in our unweighted multivariable logistic regression model (outcome adjustment model). Third, we used the stabilized mortality ratio weight, defined as 1 for immunosuppressants users and PS(1 – PS) for non-users, in place of IPTW in our main model (SMR weighting). Finally, we considered a propensity score matching approach (PS matching). All statistical analyses were conducted using R 3.5.2. P-values less than 0.05 were considered statistically significant.

## Results

4,349 patients were included in this study, for which 1,356 (31%) used immunosuppressants within 120 days of hospitalization for COVID-19. Users of immunosuppressants were older, and a larger proportion had hyperlipidemia and hypertension. A larger proportion of immunosuppressants users also used acetaminophen, systematic antibiotics, and NSAIDs (Table 1).

Patients who used immunosuppressants were at increased odds of the primary outcome of all-cause death, ICU admission, mechanical ventilation use, MI, stroke and TIA (IPTW OR 1.32; 95% CI: 1.06 – 1.63). When assessing the component outcomes individually, immunosuppressants users were only at higher odds of all-cause mortality – IPTW OR 1.63; 95% CI: 1.21 – 2.26 (Table 2).

**Table 2.** Odds ratio of adverse clinical outcomes associated with immunosuppressants use compared with non-users patients with COVID-19.

|  | Number<br>of<br>Patients | Number<br>of<br>Events | Cumulative<br>Incidence (%) | Odds ratio (95% Confidence Interval) |  |  |
| --- | --- | --- | --- | --- | --- | --- |
|  |  |  |  | Unadjusted* | Adjusted§ | IPTW Weighted# |
| **All-cause death, ICU admission, mechanical ventilation use** |  |  |  |  |  |  |
| Non-users | 2993 | 311 | 10.4 | 1.00 (reference) | 1.00 (reference) | 1.00 (reference) |
| Users | 1356 | 185 | 13.6 | 1.36 (1.12-1.66) | 1.31 (1.07-1.60) | 1.32 (1.06-1.63) |
| **All-cause death** |  |  |  |  |  |  |
| Non-users | 2993 | 126 | 4.2 | 1.00 (reference) | 1.00 (reference) | 1.00 (reference) |
| Users | 1356 | 91 | 6.7 | 1.64 (1.24-2.16) | 1.68 (1.23-2.30) | 1.63 (1.21-2.26) |
| **Mechanical ventilation** |  |  |  |  |  |  |
| Non-users | 2993 | 72 | 2.4 | 1.00 (reference) | 1.00 (reference) | 1.00 (reference) |
| Users | 1356 | 51 | 3.8 | 1.59 (1.10-2.28) | 1.48 (1.02-2.14) | 1.12 (0.76-1.65) |
| **ICU admission** |  |  |  |  |  |  |
| Non-users | 2993 | 212 | 7.1 | 1.00 (reference) | 1.00 (reference) | 1.00 (reference) |
| Users | 1356 | 118 | 8.7 | 1.25 (0.99-1.58) | 1.22 (0.96-1.54) | 1.11 (0.87-1.44) |
| **Mechanical ventilation, ICU admission** |  |  |  |  |  |  |
| Non-users | 2993 | 237 | 7.9 | 1.00 (reference) | 1.000 (reference) | 1.00 (reference) |
| Users | 1356 | 136 | 10.0 | 1.30 (1.04-1.62) | 1.25 (1.00-1.57) | 1.12 (0.88-1.42) |

\* Unweighted univariable logistic regression model
§ Unweighted multivariable logistic regression model adjusted for age, sex, insurance type, history of diabetes and history of hypertension

There exists no effect modification by age, sex, history of autoimmune diseases, cancer and HIV, and history of autoimmune disease (Figure 2). The results of the sensitivity analysis remained consistent with the main analysis (Supplementary Material 3-6). When redefining the study population to include all confirmed patients with COVID-19 (IPTW OR 1.33; 95% CI: 1.07 – 1.65), changing the exposure ascertainment window to 90 days (IPTW OR 1.43; 95% CI: 1.15 – 1.77), and applying other statistical methods (IPTW with trimming (IPTW OR 1.32; 95% CI: 1.06 – 1.63), outcome adjustment model (OR 1.29; 95% CI: 1.05 – 1.58), SMR weighting (OR 1.30; 95% CI: 1.03 – 1.64), PS matching (OR 1.32; 95% CI: 1.07 – 1.64)), immunosuppressants use was associated with an increased odd of the primary outcome consistent with the main analysis (Supplementary Material 3).

**Figure 2.**
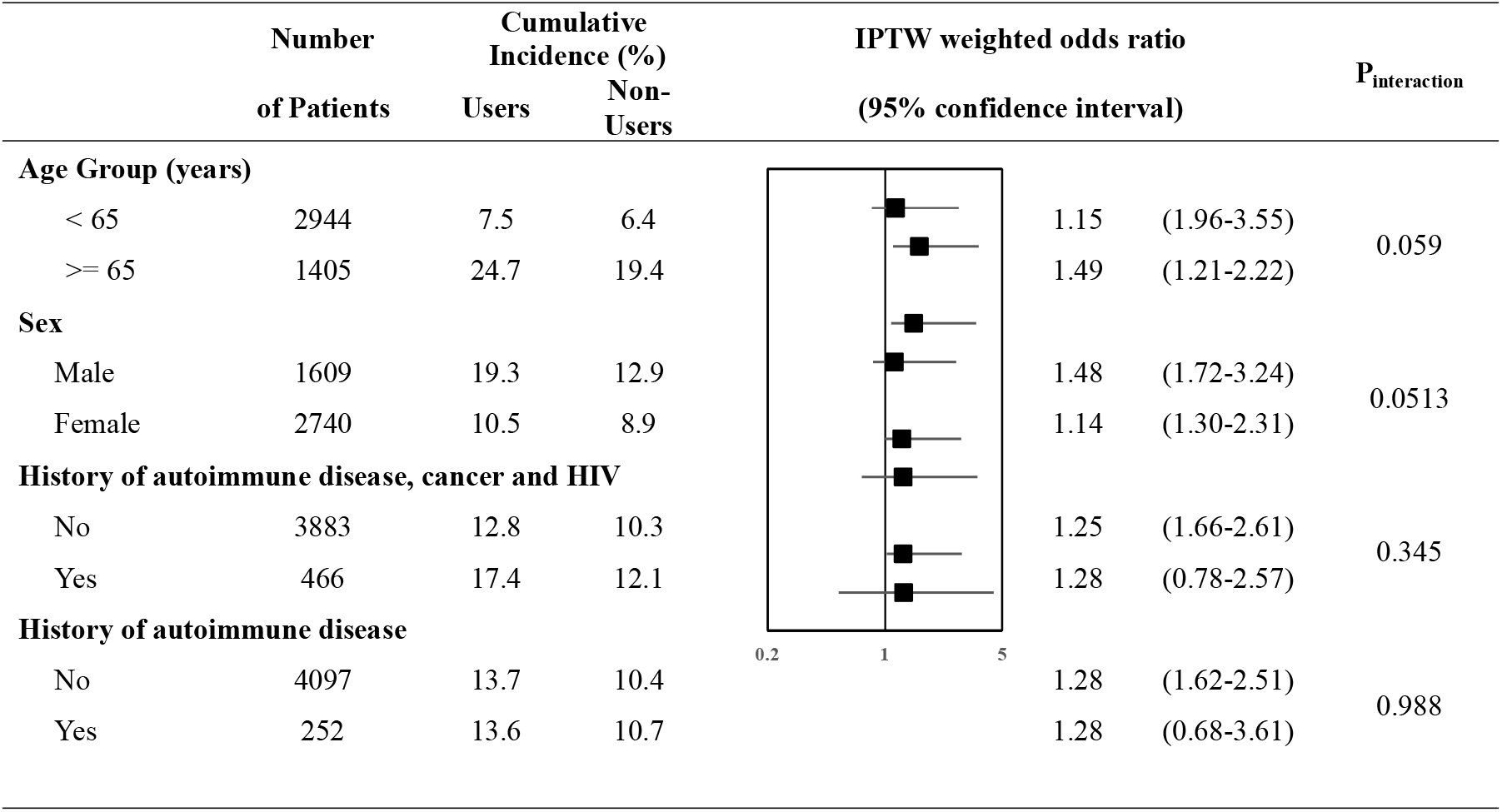
Forest plot summarizing the Odds ratio of primary outcome associated with Immunosuppressant use compared with non-users when stratified for age, sex, history of autoimmune disease, cancer and HIV, and history of autoimmune disease.

When assessing the component outcomes individually, immunosuppressants users were only at higher odds of all-cause mortality while redefining the study population to include all confirmed patients with COVID-19 (IPTW OR 1.67; 95% CI: 1.23 – 2.26), changing the exposure ascertainment window to 90 days (IPTW OR 1.85; 95% CI: 1.36 – 2.52), and applying other statistical methods (IPTW with trimming (IPTW OR 1.65; 95% CI: 1.21 – 2.26), outcome adjustment model (OR 1.59; 95% CI: 1.18 – 2.13), SMR weighting (OR 1.51; 95% CI: 1.05 – 2.16), PS matching (OR 1.57; 95% CI: 1.14 – 2.17)).

Among 1,356 immunosuppressants users in the main analysis, 1,340 (98.8%) were corticosteroid users. They were at increased odds of the primary outcome (IPTW OR 1.33; 95% CI: 1.07 – 1.64) (Supplementary Material 3). When assessing the component outcomes individually, corticosteroids users were only at higher odds of all-cause mortality – IPTW OR 1.67; 95% CI: 1.22 – 2.76 (Supplementary Material 4-6), which is consistent with the analysis for immunosuppressants use.

## Discussion

This study reports on a nationwide cohort of South Korean COVID-19 patients. This dataset was completely enumerated, and statistically controlled for confounding using propensity scores. The results suggest that patients administered immunosuppressants were at increased odds of all-cause mortality, mechanical ventilation and ICU admission,

Our results supports the guideline from the Center for Disease Control and Prevention in the United States, classifying patients with prior use of corticosteroids and other immuosuppressive medication as a high risk group of patients for severe COVID-19^12^. In support of this assertion are papers by Brenner *et al*^13^, Michelena *et al*^14^, Di Giorgio *et al*^15^, Marlais *et al*^16^ and Montero-Escribano *et al*^17^.

Corticosteroids are known to reduce mortality, mechanical ventilation and duration of hospital stay relative to standard-of-care in patients with severe and critical COVID-19, as noted by the living systematic review and meta-analysis by Siemieniuk *et al*^18^ and WHO clinical guideline^7^.

Also, Anderson *et al*^19^ reported contrary results to our paper, in that adverse outcomes were not associated with immunosuppressants use before COVID-19. However, their paper reported on a sicker patient population who might have received benefit from corticosteroids, whereas our analysis reports on a nationwide cohort of all patients admitted for COVID-19 to South Korean hospitals which include patients with non-severe COVID-19 infection. In South Korea, typically all COVID-19 patients were admitted to hospital even if they were asymptomatic or had mild symptoms, in an attempt to limit spread of the COVID-19 infection. As a result, our study provides a more complete picture with all spectrum of COVID-19 patients, from asymptomatic to critical COVID-19. As corticosteroids use is beneficial in patients with severe to critical stage in COVID-19 infection and could possibly be harmful in patients with non-severe COVID-19 infection, as seen from RECOVERY trial^8^, it comes as no surprise that our study results showing increased odds of primary outcome or mortality in immunosuppressants or corticosteroids user given that most of COVID-19 infections are non-severe^20^.

Given our study results, we support booster COVID-19 vaccinations for immunocompromised people and urge caution around blanket-continuation of immunosuppressants for COVID-19 who were on immunosuppressants prior to COVID-19 diagnosis; patients might need to be assessed on a per-case basis, weighting the risk of adverse COVID-19 outcomes and benefits of continuing immunosuppressants. Consideration may need to be given to lower the degree of immunosuppression if a patient was on corticosteroids and has non-severe COVID-19 infection.

There are several notable strengths of this study. As South Korea used a strict nationwide patient management system for COVID-19, the use of a population-based cohort mitigated any potential sampling bias issue. There could be very little chance of outcome misclassification since outcomes such as all-cause death and outcomes defined from procedures codes (i.e. ICU admissions and mechanical ventilation use) are unlikely to be misclassified in the main analysis restricted to hospitalized COVID-19 patients, given the cross-referencing of these records with national death records and reimbursement review processes, respectively. Consistent results across rigorous statistical methods including IPTW, PS matching, SMR and thorough sensitivity analyses suggest a robustness of our result. Additionally, including chronic co-medications as confounders could have mitigated healthy user bias.

This study was also not without limitations. We do not have data regarding the severity of COVID-19 at the time of COVID-19 diagnosis, which is a limitation of using claims data. In addition, there may still exist residual confounding by confounders that are typically not captured in a claims database (e.g., body mass index, baseline blood pressure, laboratory test values). Also, our result is limited by the observational nature of our study design.

In conclusion, our study of a large nationwide cohort of hospitalized COVID-19 patients in South Korea finds that use of immunosuppressants or corticosteroids increases the odds of all-cause mortality, mechanical ventilation and ICU admissions. We support the latest guidelines from the CDC, that people on immunosuppressants are at high risk of severe COVID-19 and immunocompromised people need booster COVID-19 vaccinations.

## Acknowledgements

The authors thank healthcare professionals dedicated to treating COVID-19 patients in South Korea, the Ministry of Health and Welfare, the Health Insurance Review & Assessment Service (HIRA) and Ye-Jin Sohn (HIRA) for sharing invaluable national health insurance claims data.

## Data sharing

No additional data available.

## Supplementary Material Legends

**Supplementary Material 1**. Data schema of the Health Insurance Review and Assessment Service database.

### 1. Data Description

The database used in this study is the Health Insurance Review and Assessment Service (HIRA) claim data released by the South Korean government as the world’s first de-identified COVID-19 nationwide data. Since South Korea have implemented a single National Health Insurance system, this administrative data includes the entire health insurance claims and clinical information of South Korean population. Information on non-essential medical treatment that are not covered by NHI was not collected based on fee-for-service payment system.

### 2. Data Scope

The list of patients of COVID-19 (using submitted claims data) is connected to their history of medical service use for the past 5 years (using finalized claims data, from January 2015 to February 2020), and the entire data set is de-identified. All benefit claims relevant to COVID-19 as of May 15, 2020 were included.

### 3. Data Extraction Criteria

The databased include all health insurance types such as National Health Insurance, Medical Aid, Korea veterans service. COVID19 related claims consists of i) claims with “3/02” in classification code of MT043 (Target of medical cost support due to national disaster) based on the serial number of claim statement, ii) claims that contained “D6584” COVID-19 (Real-time polymerase chain reaction), iii) claims that contained COVID-19 related disease code (B342, B972, Z208, Z290, U18, U181, Z038, Z115, U071, U072), and iv)□claims that contained other COVID-19 related fee code (COVID-19 related admission fee, management fee, IUD, isolation, public relief hospital, residential treatment center, screening center, negative pressure room, etc.).

### 4. Data Schema

The data based consists of COVID-19 related claims data, 3years of healthcare service use history of the individuals who submitted COVID-19 related claims. COVID-19 confirmation information and some related results were extracted from KCDA database. The included information are as follows:

- Sociodemographic characteristics (i.e., sex, age, medical institution type)
- Diagnosis information (i.e., ICD-10 diagnosis codes, main diagnosis, sub diagnosis)
- Healthcare utilization information (i.e., number of visit, date of visit, duration of hospital stay)
- Prescription information (i.e., national drug chemical code, number of supply, date of supply, dosage)

**Note**: COVID-19, coronavirus disease 2019; ICD-10, International Classification of Disease 10^th^ Revision. Detailed information on data schema is available upon request.

**Supplementary Material 2**. Diagnosis codes based on the Korean Standard Classification of Diseases, 7^th^ Revision or International Classification of Disease, 10^th^ Revision codes, National Procedure codes, and drug codes based on World Health Organization-Anatomical Therapeutic Chemical classification codes

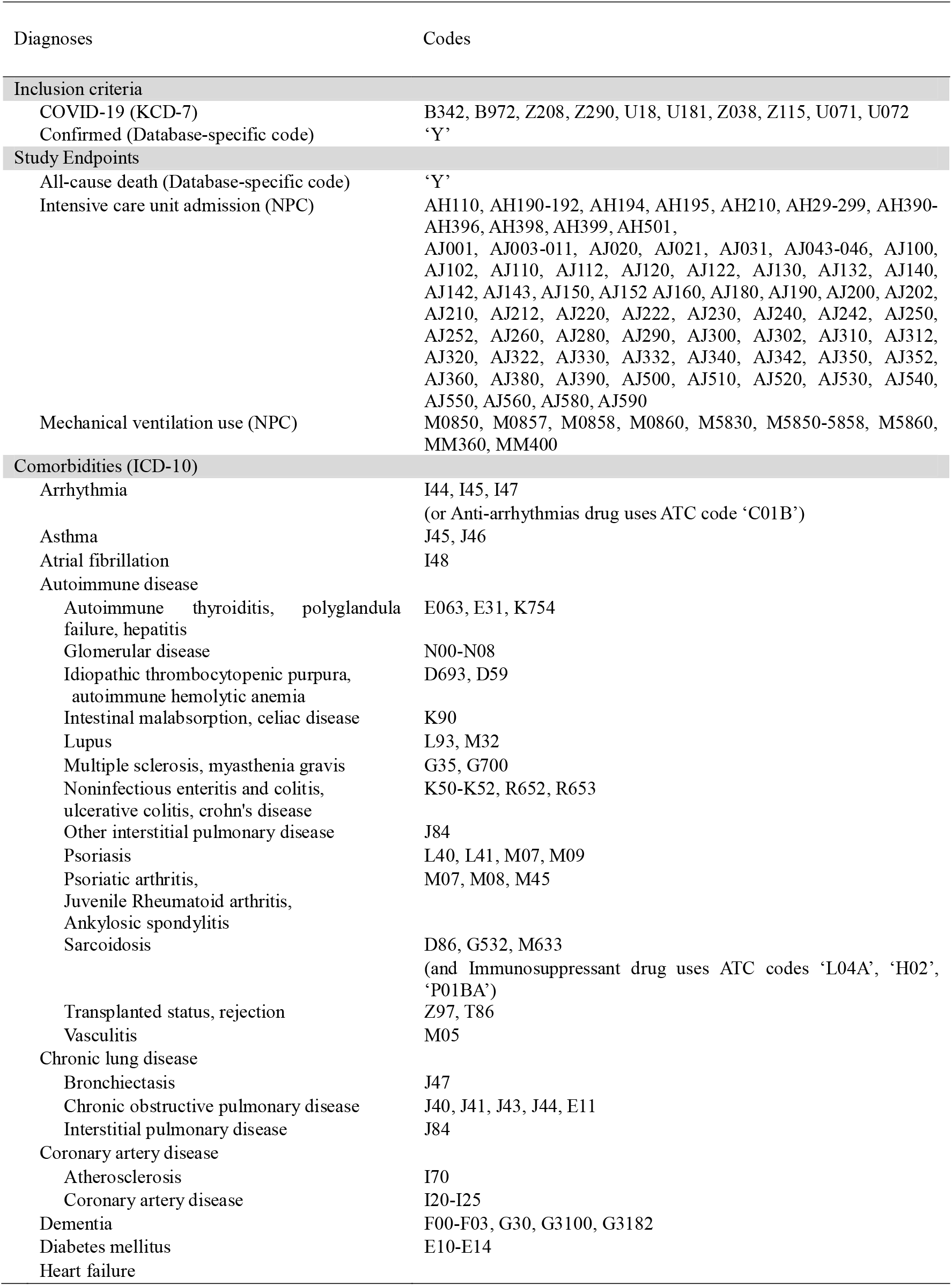
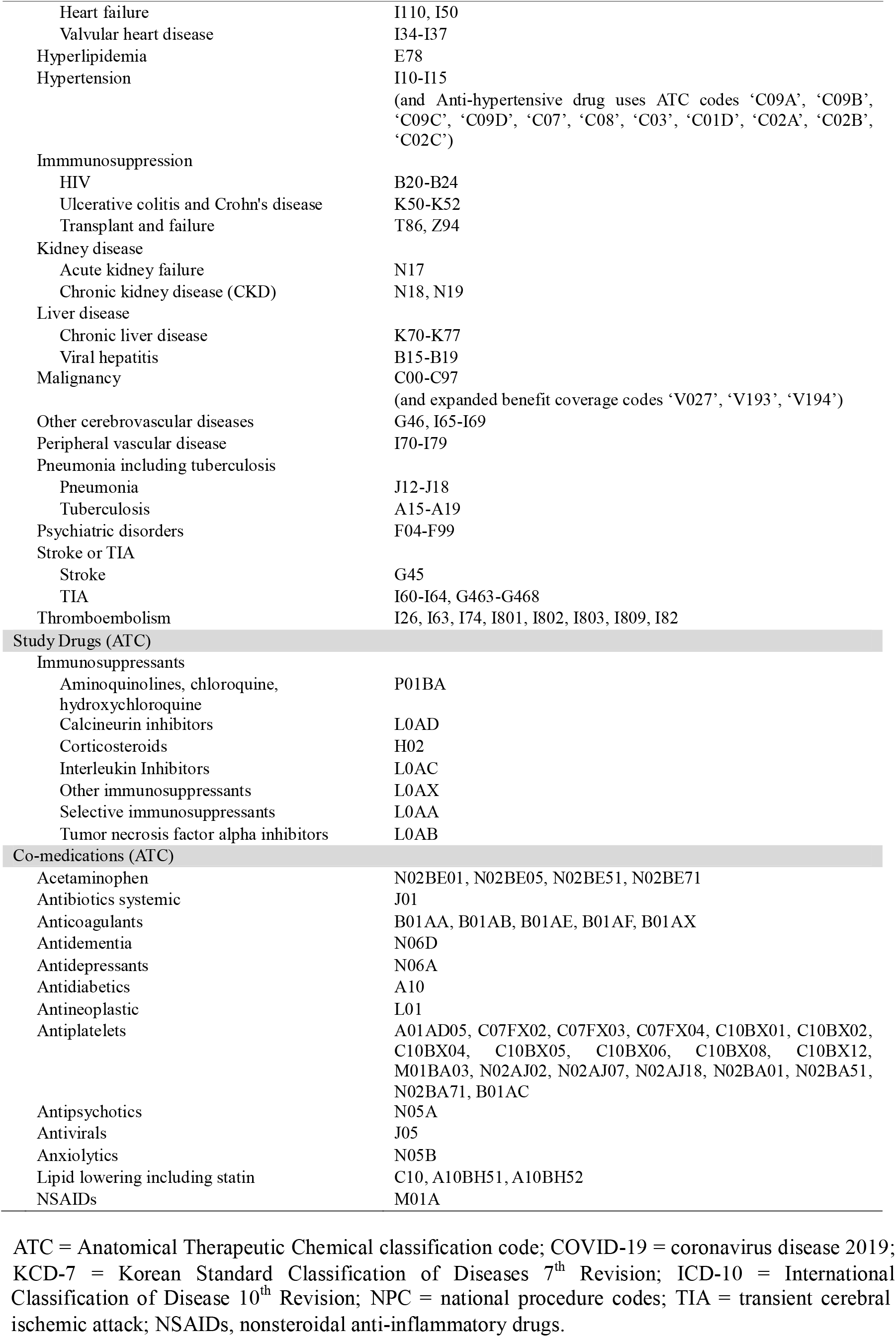

| Diagnoses | Codes |
| --- | --- |
| **Inclusion criteria** |  |
| COVID-19 (KCD-7) | B342, B972, Z208, Z290, U18, U181, Z038, Z115, U071, U072 |
| Confirmed (Database-specific code) | ‘Y’ |
| **Study Endpoints** |  |
| All-cause death (Database-specific code) | ‘Y’ |
| Intensive care unit admission (NPC) | AH110, AH190-192, AH194, AH195, AH210, AH29-299, AH390-AH396, AH398, AH399, AH501, AJ001, AJ003-011, AJ020, AJ021, AJ031, AJ043-046, AJ100, AJ102, AJ110, AJ112, AJ120, AJ122, AJ130, AJ132, AJ140, AJ142, AJ143, AJ150, AJ152 AJ160, AJ180, AJ190, AJ200, AJ202, AJ210, AJ212, AJ220, AJ222, AJ230, AJ240, AJ242, AJ250, AJ252, AJ260, AJ280, AJ290, AJ300, AJ302, AJ310, AJ312, AJ320, AJ322, AJ330, AJ332, AJ340, AJ342, AJ350, AJ352, AJ360, AJ380, AJ390, AJ500, AJ510, AJ520, AJ530, AJ540, AJ550, AJ560, AJ580, AJ590 |
| Mechanical ventilation use (NPC) | M0850, M0857, M0858, M0860, M5830, M5850-5858, M5860, MM360, MM400 |
| **Comorbidities (ICD-10)** |  |
| Arrhythmia | I44, I45, I47<br>(or Anti-arrhythmias drug uses ATC code ‘C01B’) |
| Asthma | J45, J46 |
| Atrial fibrillation | I48 |
| Autoimmune disease |  |
| Autoimmune thyroiditis, polyglandula failure, hepatitis | E063, E31, K754 |
| Glomerular disease | N00-N08 |
| Idiopathic thrombocytopenic purpura, autoimmune hemolytic anemia | D693, D59 |
| Intestinal malabsorption, celiac disease | K90 |
| Lupus | L93, M32 |
| Multiple sclerosis, myasthenia gravis | G35, G700 |
| Noninfectious enteritis and colitis, ulcerative colitis, crohn's disease | K50-K52, R652, R653 |
| Other interstitial pulmonary disease | J84 |
| Psoriasis | L40, L41, M07, M09 |
| Psoriatic arthritis, Juvenile Rheumatoid arthritis, Ankylosic spondylitis | M07, M08, M45 |
| Sarcoidosis | D86, G532, M633<br>(and Immunosuppressant drug uses ATC codes ‘L04A’, ‘H02’, ‘P01BA’) |
| Transplanted status, rejection | Z97, T86 |
| Vasculitis | M05 |
| Chronic lung disease |  |
| Bronchiectasis | J47 |
| Chronic obstructive pulmonary disease | J40, J41, J43, J44, E11 |
| Interstitial pulmonary disease | J84 |
| Coronary artery disease |  |
| Atherosclerosis | I70 |
| Coronary artery disease | I20-I25 |
| Dementia | F00-F03, G30, G3100, G3182 |
| Diabetes mellitus | E10-E14 |
| Heart failure |  |
| Heart failure | I110, I50 |
| Valvular heart disease | I34-I37 |
| Hyperlipidemia | E78 |
| Hypertension | I10-I15<br>(and Anti-hypertensive drug uses ATC codes 'C09A', 'C09B', 'C09C', 'C09D', 'C07', 'C08', 'C03', 'C01D', 'C02A', 'C02B', 'C02C') |
| Immunosuppression |  |
| HIV | B20-B24 |
| Ulcerative colitis and Crohn's disease | K50-K52 |
| Transplant and failure | T86, Z94 |
| Kidney disease |  |
| Acute kidney failure | N17 |
| Chronic kidney disease (CKD) | N18, N19 |
| Liver disease |  |
| Chronic liver disease | K70-K77 |
| Viral hepatitis | B15-B19 |
| Malignancy | C00-C97<br>(and expanded benefit coverage codes 'V027', 'V193', 'V194') |
| Other cerebrovascular diseases | G46, I65-I69 |
| Peripheral vascular disease | I70-I79 |
| Pneumonia including tuberculosis |  |
| Pneumonia | J12-J18 |
| Tuberculosis | A15-A19 |
| Psychiatric disorders | F04-F99 |
| Stroke or TIA |  |
| Stroke | G45 |
| TIA | I60-I64, G463-G468 |
| Thromboembolism | I26, I63, I74, I801, I802, I803, I809, I82 |
| **Study Drugs (ATC)** |  |
| Immunosuppressants |  |
| Aminoquinolines, chloroquine, hydroxychloroquine | P01BA |
| Calcineurin inhibitors | L0AD |
| Corticosteroids | H02 |
| Interleukin Inhibitors | L0AC |
| Other immunosuppressants | L0AX |
| Selective immunosuppressants | L0AA |
| Tumor necrosis factor alpha inhibitors | L0AB |
| **Co-medications (ATC)** |  |
| Acetaminophen | N02BE01, N02BE05, N02BE51, N02BE71 |
| Antibiotics systemic | J01 |
| Anticoagulants | B01AA, B01AB, B01AE, B01AF, B01AX |
| Antidementia | N06D |
| Antidepressants | N06A |
| Antidiabetics | A10 |
| Antineoplastic | L01 |
| Antiplatelets | A01AD05, C07FX02, C07FX03, C07FX04, C10BX01, C10BX02, C10BX04, C10BX05, C10BX06, C10BX08, C10BX12, M01BA03, N02AJ02, N02AJ07, N02AJ18, N02BA01, N02BA51, N02BA71, B01AC |
| Antipsychotics | N05A |
| Antivirals | J05 |
| Anxiolytics | N05B |
| Lipid lowering including statin | C10, A10BH51, A10BH52 |
| NSAIDs | M01A |

ATC = Anatomical Therapeutic Chemical classification code; COVID-19 = coronavirus disease 2019; KCD-7 = Korean Standard Classification of Diseases 7<sup>th</sup> Revision; ICD-10 = International
Classification of Disease 10<sup>th</sup> Revision; NPC = national procedure codes; TIA = transient cerebral ischemic attack; NSAIDs, nonsteroidal anti-inflammatory drugs.

**Supplementary Material 3**. Odds ratio of primary outcome associated with immunosuppressants use compared with non-use among COVID-19 patients with >= 40 years of age.

|  | Number<br>of Patients | Cumulative<br>Incidence (%)<br>Non-Users | Users |  | Odds<br>Ratio | (95%<br>Confidence<br>interval) |
| --- | --- | --- | --- | --- | --- | --- |
| All-cause Death, ICU admission, Mechanical ventilation use |  |  |  |  |  |  |
| Confirmed-patients with COVID-19 |  |  |  |  |  |  |
| Immunosuppressant | 4613 | 10.0 | 13.3 |  | 1.33 | (1.07-1.65) |
| Corticosteroids | 4597 | 10.0 | 13.5 |  | 1.34 | (1.08-1.65) |
| Redefining the exposure searching window |  |  |  |  |  |  |
| 90 days before and including the date of cohort entry |  |  |  |  |  |  |
| Immunosuppressant | 4613 | 9.9 | 14.2 |  | 1.43 | (1.15-1.77) |
| Corticosteroids | 4598 | 9.9 | 14.3 |  | 1.44 | (1.16-1.78) |
| Hospitalized-patients with COVID-19 |  |  |  |  |  |  |
| Immunosuppressant | 4349 | 10.4 | 13.6 |  | 1.32 | (1.06-1.63) |
| Corticosteroids | 4333 | 10.4 | 13.8 |  | 1.33 | (1.07-1.64) |
| Redefining the exposure searching window |  |  |  |  |  |  |
| 90 days before and including the date of cohort entry |  |  |  |  |  |  |
| Immunosuppressant | 4349 | 10.3 | 14.6 |  | 1.43 | (1.15-1.77) |
| Corticosteroids | 4334 | 10.3 | 14.7 |  | 1.44 | (1.16-1.79) |
| Statistical Method based on Propensity Score (Immunosuppressant) |  |  |  |  |  |  |
| IPTW with trimming | 4349 | 10.4 | 13.6 |  | 1.32 | (1.06-1.63) |
| Outcome Adjustment model | 4349 | 10.4 | 13.6 |  | 1.29 | (1.05-1.58) |
| SMR weighting | 4349 | 10.4 | 13.6 |  | 1.30 | (1.03-1.64) |
| PS Matching | 4349 | 10.4 | 13.6 |  | 1.32 | (1.07-1.64) |

**Supplementary Material 4**. Odds ratio of all-cause death associated with immunosuppressants use compared with non-use among COVID-19 patients with >= 40 years of age.

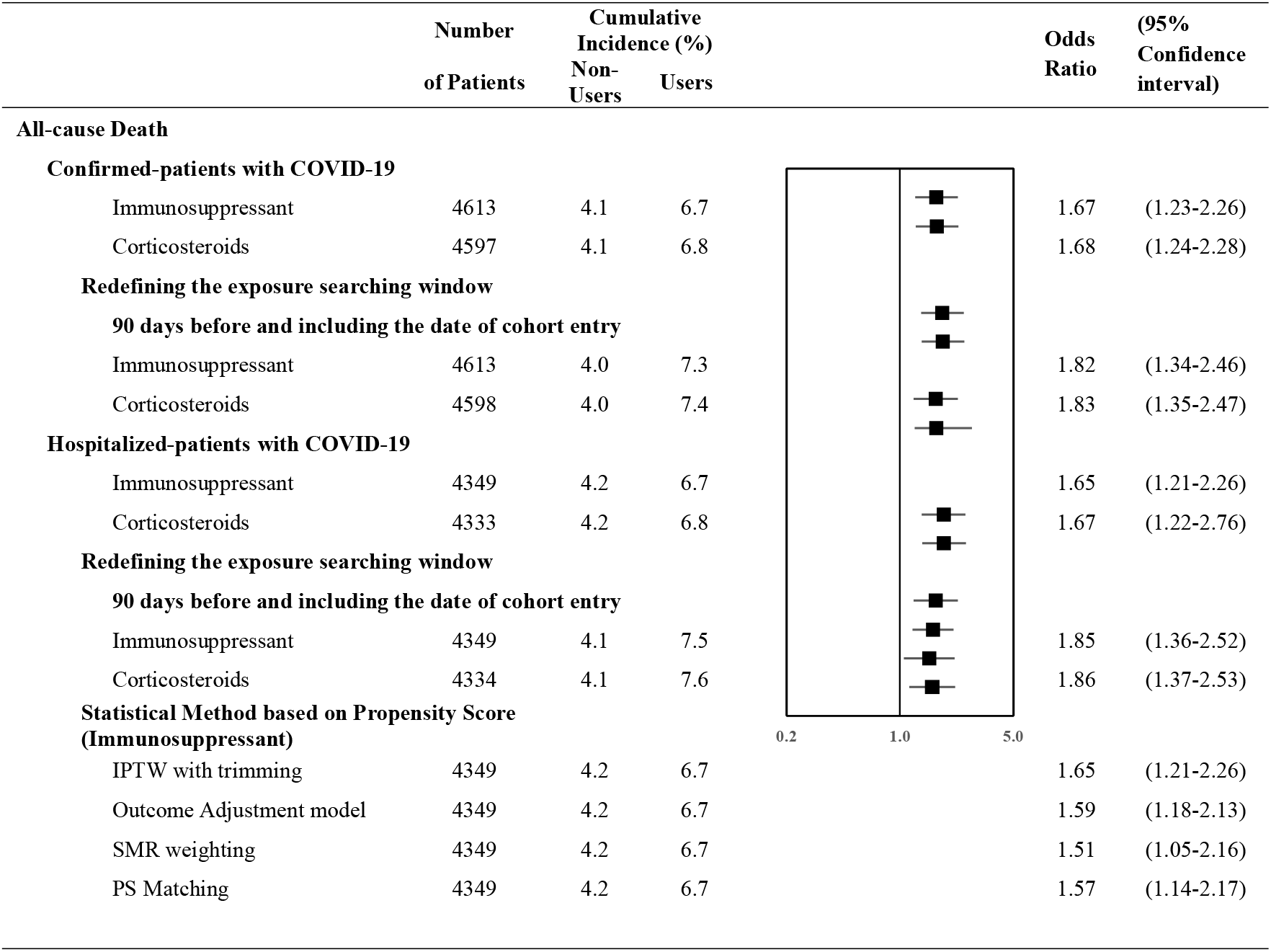

**Supplementary Material 5**. Odds ratio of adverse mechanical ventilation associated with immunosuppressants use compared with non-use among COVID-19 patients with >= 40 years of age.

|  | Number<br>of Patients | Cumulative<br>Incidence (%)<br>Non-Users | Users |  | Odds<br>Ratio | (95%<br>Confidence<br>interval) |
| --- | --- | --- | --- | --- | --- | --- |
| **Mechanical ventilation** |  |  |  |  |  |  |
| **Confirmed-patients with COVID-19** |  |  |  |  |  |  |
| Immunosuppressant | 4613 | 2.3 | 3.6 |  | 1.13 | (0.77-1.66) |
| Corticosteroids | 4597 | 2.3 | 3.6 |  | 1.13 | (0.77-1.66) |
| **Redefining the exposure searching window** |  |  |  |  |  |  |
| **90 days before and including the date of cohort entry** |  |  |  |  |  |  |
| Immunosuppressant | 4613 | 2.2 | 3.9 |  | 1.37 | (0.92-2.02) |
| Corticosteroids | 4598 | 2.2 | 3.9 |  | 1.37 | (0.93-2.02) |
| **Hospitalized-patients with COVID-19** |  |  |  |  |  |  |
| Immunosuppressant | 4349 | 2.4 | 3.8 |  | 1.12 | (0.76-1.65) |
| Corticosteroids | 4333 | 2.4 | 3.8 |  | 1.13 | (0.77-1.65) |
| **Redefining the exposure searching window** |  |  |  |  |  |  |
| **90 days before and including the date of cohort entry** |  |  |  |  |  |  |
| Immunosuppressant | 4349 | 2.4 | 4.1 |  | 1.36 | (0.92-2.00) |
| Corticosteroids | 4334 | 2.4 | 4.1 |  | 1.36 | (0.92-2.01) |
| **Statistical Method based on Propensity Score (Immunosuppressant)** |  |  |  |  |  |  |
| IPTW with trimming | 4349 | 2.4 | 3.8 |  | 1.12 | (0.76-1.65) |
| Outcome Adjustment model | 4349 | 2.4 | 3.8 |  | 1.20 | (0.81-1.77) |
| SMR weighting | 4349 | 2.4 | 3.8 |  | 1.27 | (0.83-1.92) |
| PS Matching | 4349 | 2.4 | 3.8 |  | 1.27 | (0.84-1.92) |

**Supplementary Material 6**. Odds ratio of ICU admission associated with immunosuppressants use among COVID-19 patients with >=40 years of age.

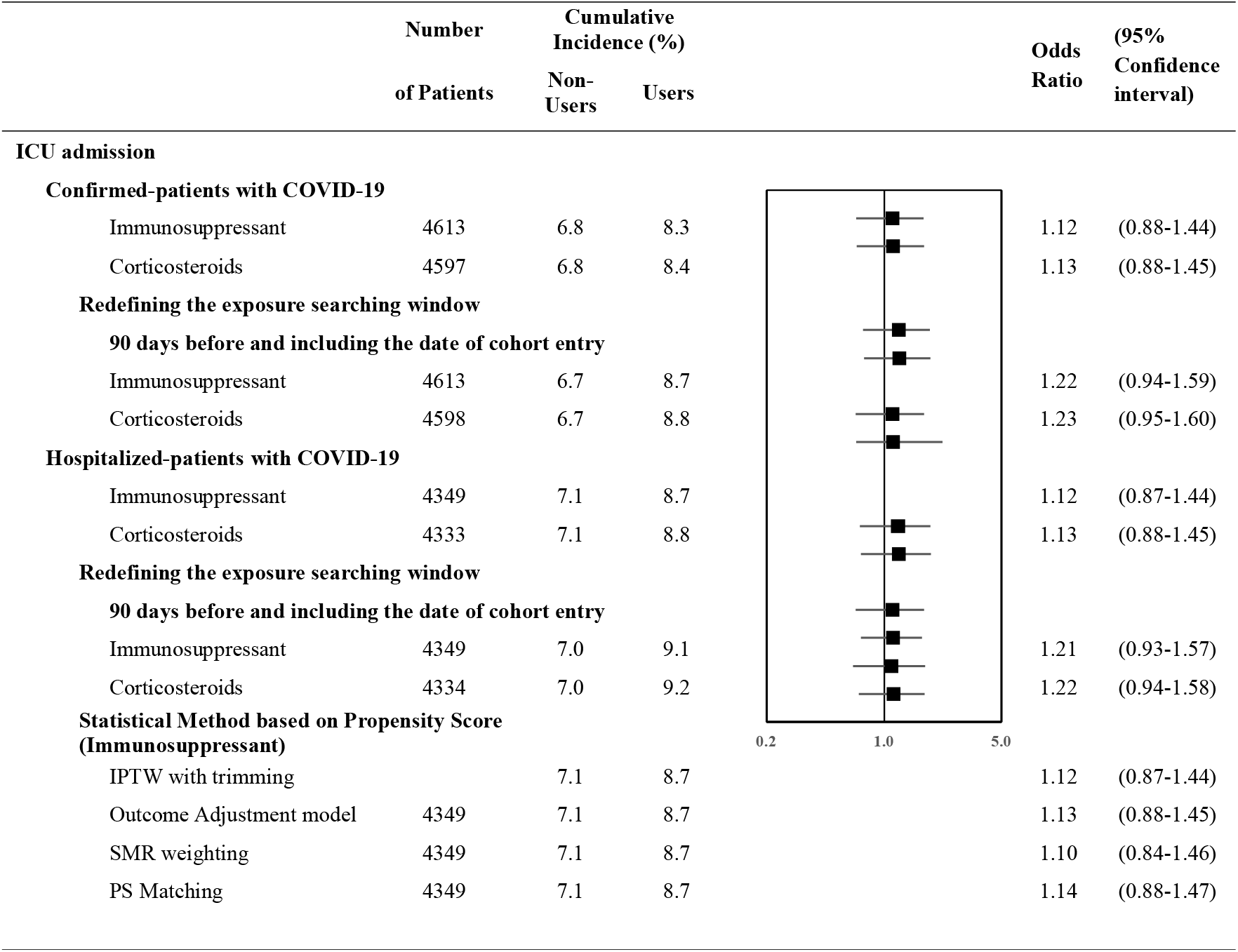

|  | Number | Cumulative Incidence (%) |  |  | Odds Ratio | (95% Confidence interval) |
| --- | --- | --- | --- | --- | --- | --- |
|  | of Patients | Non-Users | Users |  |  |  |
| **ICU admission** |  |  |  |  |  |  |
| **Confirmed-patients with COVID-19** |  |  |  |  |  |  |
| Immunosuppressant | 4613 | 6.8 | 8.3 |  | 1.12 | (0.88-1.44) |
| Corticosteroids | 4597 | 6.8 | 8.4 |  | 1.13 | (0.88-1.45) |
| **Redefining the exposure searching window** |  |  |  |  |  |  |
| **90 days before and including the date of cohort entry** |  |  |  |  |  |  |
| Immunosuppressant | 4613 | 6.7 | 8.7 |  | 1.22 | (0.94-1.59) |
| Corticosteroids | 4598 | 6.7 | 8.8 |  | 1.23 | (0.95-1.60) |
| **Hospitalized-patients with COVID-19** |  |  |  |  |  |  |
| Immunosuppressant | 4349 | 7.1 | 8.7 |  | 1.12 | (0.87-1.44) |
| Corticosteroids | 4333 | 7.1 | 8.8 |  | 1.13 | (0.88-1.45) |
| **Redefining the exposure searching window** |  |  |  |  |  |  |
| **90 days before and including the date of cohort entry** |  |  |  |  |  |  |
| Immunosuppressant | 4349 | 7.0 | 9.1 |  | 1.21 | (0.93-1.57) |
| Corticosteroids | 4334 | 7.0 | 9.2 |  | 1.22 | (0.94-1.58) |
| **Statistical Method based on Propensity Score (Immunosuppressant)** |  |  |  |  |  |  |
| IPTW with trimming |  | 7.1 | 8.7 |  | 1.12 | (0.87-1.44) |
| Outcome Adjustment model | 4349 | 7.1 | 8.7 |  | 1.13 | (0.88-1.45) |
| SMR weighting | 4349 | 7.1 | 8.7 |  | 1.10 | (0.84-1.46) |
| PS Matching | 4349 | 7.1 | 8.7 |  | 1.14 | (0.88-1.47) |

## References

1. Lai CC, Shih TP, Ko WC, Tang HJ, Hsueh PR. Severe acute respiratory syndrome coronavirus 2 (SARS-CoV-2) and coronavirus disease-2019 (COVID-19): The epidemic and the challenges. Int J Antimicrob Agents 2020; 55(3): 105924.

2. Gorbalenya AE, Baker SC, Baric R, et al. Severe acute respiratory syndrome-related coronavirus: The species and its viruses – a statement of the Coronavirus Study Group. bioRxiv 2020.

3. Chow R, Simone CB, 2nd, Lock M. Hydroxychloroquine for the treatment of COVID-19: the importance of scrutiny of positive trials. Ann Palliat Med 2020.

4. Baden LR, Rubin EJ. Covid-19 - The Search for Effective Therapy. N Engl J Med 2020; 382(19): 1851–2.

5. Mehta P, McAuley DF, Brown M, Sanchez E, Tattersall RS, Manson JJ. COVID-19: consider cytokine storm syndromes and immunosuppression. The Lancet 2020; 395(10229): 1033–4.

6. Russell CD, Millar JE, Baillie JK. Clinical evidence does not support corticosteroid treatment for 2019-nCoV lung injury. The Lancet 2020; 395(10223): 473–5.

7. Update to living WHO guideline on drugs for covid-19. Bmj 2020; 371: m4475.

8. Horby P, Lim WS, Emberson JR, et al. Dexamethasone in Hospitalized Patients with Covid-19. N Engl J Med 2021; 384(8): 693–704.

9. Republic of Korea Ministry of Health and Welfare. COVID-19 Response: Korean Government’s Response System. 2021. http://ncov.mohw.go.kr/en/baroView.do?brdId=11&brdGubun=111 (Accessed 1 August 2021.

10. Freemantle N, Marston L Fau - Walters K, Walters K Fau - Wood J, Wood J Fau - Reynolds MR, Reynolds Mr Fau - Petersen I, Petersen I. Making inferences on treatment effects from real world data: propensity scores, confounding by indication, and other perils for the unwary in observational research. BMJ 2013; 347: f6409.

11. Imbens GW, Rubin DB. Causal Inference for Statistics, Social, and Biomedical Sciences: Cambridge University Press; 2015.

12. Centers for Disease Control and Prevention. Underlying medical conditions associated with high risk for severe COVID-19: information for healthcare providers. 2021. https://www.cdc.gov/coronavirus/2019-ncov/hcp/clinical-care/underlyingconditions.html (Accessed 15 June 2021.

13. Brenner EJ, Ungaro RC, Gearry RB, et al. Corticosteroids, But Not TNF Antagonists, Are Associated With Adverse COVID-19 Outcomes in Patients With Inflammatory Bowel Diseases: Results From an International Registry. Gastroenterology 2020; 159(2): 481-91.e3.

14. Michelena X, Borrell H, López-Corbeto M, et al. Incidence of COVID-19 in a cohort of adult and paediatric patients with rheumatic diseases treated with targeted biologic and synthetic disease-modifying anti-rheumatic drugs. Semin Arthritis Rheum 2020; 50(4): 564–670.

15. Di Giorgio A, Nicastro E, Speziani C, et al. Health status of patients with autoimmune liver disease during SARS-CoV-2 outbreak in northern Italy. Journal of Hepatology 2020; 73(3): 702–5.

16. Marlais M, Wlodkowski T, Vivarelli M, et al. The severity of COVID-19 in children on immunosuppressive medication. The Lancet Child & Adolescent Health 2020; 4(7): e17–e8.

17. Montero-Escribano P, Matías-Guiu J, Gómez-Iglesias P, Porta-Etessam J, Pytel V, Matias-Guiu JA. Anti-CD20 and COVID-19 in multiple sclerosis and related disorders: A case series of 60 patients from Madrid, Spain. Multiple Sclerosis and Related Disorders 2020; 42.

18. Siemieniuk RA, Bartoszko JJ, Ge L, et al. Drug treatments for covid-19: living systematic review and network meta-analysis. Bmj 2020; 370: m2980.

19. Andersen KM, Mehta HB, Palamuttam N, et al. Association Between Chronic Use of Immunosuppresive Drugs and Clinical Outcomes From Coronavirus Disease 2019 (COVID-19) Hospitalization: A Retrospective Cohort Study in a Large US Health System. Clin Infect Dis 2021.

20. Kim AY, Gandhi RT. COVID-19: Management in hospitalized adults. 2021. https://www.uptodate.com/contents/covid-19-management-in-hospitalized-adults/print?search=statincovid&source=search_result&selectedTitle=1~150&usage_type=default&display_rank=11/37 (Accessed 10 July 2021).

